# COVID-19 vaccine acceptance in older Syrian refugees: preliminary findings from an ongoing study

**DOI:** 10.1101/2021.04.25.21256024

**Authors:** Noura Salibi, Sawsan Abdulrahim, Maria El Haddad, Stephanie Bassil, Zeina El Khoury, Hala Ghattas, Stephen J. McCall

## Abstract

This study assesses COVID-19 vaccine intentions among a sample of Syrian refugees (≥50 years) beneficiaries of a humanitarian organization in Lebanon, and explores factors associated with vaccine refusal. The findings are part of an ongoing rotating 4-wave panel study. The sample was limited to participants from the first panel who completed a phone interview between January-February, 2021. Out 1,037 beneficiaries, almost a third (29%) reported no intention to vaccinate. Reasons for refusal were: newness of the vaccine (35%); preference to maintain precaution measures (21%); belief that COVID-19 vaccine is not essential (21%); and other reasons (23%). COVID-19 vaccine refusal was significantly associated with perceptions regarding vaccine safety (OR: 5.97; 95%CI: 4.03-8.84) and effectiveness (OR: 6.80; 95%CI:4.44-10.42) but did not differ by age, presence of chronic conditions, self-reported adherence to COVID-19 measures, and perceptions of susceptibility to and severity of COVID-19. Addressing vaccine hesitancy among Syrian refugees in Lebanon necessitates disseminating accurate, accessible, and culturally appropriate information about vaccine safety and effectiveness.

## Introduction

Achieving COVID-19 herd immunity worldwide demands the implementation of equitable vaccination campaigns and overcoming multiple resource constraints such as accessibility to the vaccine, logistical capacities, and vaccine hesitancy^1,2^. Existing studies on COVID-19 vaccine hesitancy have focused on adults from the general population and healthcare workers^3^, with little evidence published on the perceptions of immigrants and refugees. As displaced populations are disproportionately affected by COVID-19 due to the syndemic effects of health and social vulnerabilities^4^, including them in national vaccination campaigns is paramount. The Arab region, which hosts one of the largest refugee populations worldwide, reports very low COVID-19 vaccine acceptance rates^3^, with only 28.4% of adults in Jordan and 44% in Lebanon expressing willingness to get vaccinated^5,6^. Lebanon launched its COVID-19 vaccination campaign on February 14, 2021 pledging for an inclusive and equitable vaccination strategy. As the campaign rolls out, understanding the factors associated with vaccine acceptance and refusal is key for public health planning to achieve herd immunity.

## Materials and Methods

We present preliminary findings on COVID-19 vaccine intentions among older Syrian refugees in Lebanon. As of December 2020, Syrian refugees numbered 865,000 in a country of six million (UNHCR)^7^. The findings come from an ongoing rotating 4-wave panel study aiming to track older Syrian refugees’ vulnerability to COVID-19 and adherence to public health measures. The sampling frame for the study was a beneficiaries list of a humanitarian organization and included a probability sample of Syrian households with at least one adult aged 50 years or older. Data for the study were collected through phone interviews. The study sample size is 3,838; however, the findings presented here are limited to a sample of 1,037 from the first panel who were interviewed between January 21, 2021 and February 22, 2021.

This study was granted approval by the Social and Behavioral Sciences Institutional Research Board. Participants voluntary agreed to participate in the study following oral consent and had the right to refuse or withdraw participation at any time without affecting their access to benefits. Intention to vaccinate against COVID-19 was assessed using the following question: “If a safe and effective vaccine for COVID-19 became available, free, would you take it?” (Yes, No, Don’t know, Refuse to answer). Using unadjusted logistic regression, we examined the association between vaccine refusal and five constellations of factors: 1. Socio-demographics; 2. Presence of chronic illness; 3. General perceptions of vaccine safety and effectiveness; 4. Adherence with COVID-19 measures; 5. Perceptions of susceptibility to and severity of COVID-19.

## Results

The majority of the sample (66%) reported an intention to receive the COVID-19 vaccine if it is safe and free; almost one third (28.8%) reported no intention to vaccinate and 5.2% didn’t know or refused to answer. Among those who reported no intention to vaccinate, the reasons provided were: newness of the vaccine and wanting to wait to know more (35%); preference to maintain public health precautions (21%); belief that the vaccine is not essential (21%); and other reasons including worry about the side effects of the vaccine, interactions with other medications, and lack of trust in the system (23%).

The findings in Table 1 demonstrate that COVID-19 vaccine acceptance was not positively associated with increased age or lower educational attainment; however, odds of refusal were statistically significantly higher among refugees living outside informal tented settlements (ITS) compared to those inside ITS. Those in the 70+ age category and those reporting chronic conditions did not significantly differ from younger participants or those without chronic conditions in their vaccine acceptance. This finding has important implications given that these are the two most at risk groups of COVID-19 severe illness and mortality^8^. Perceptions of vaccine safety and effectiveness were significantly associated with vaccine acceptance; for example those who disagreed with the statement that vaccines are safe were six times more likely to report refusal to vaccinate compared to those who believed in vaccines’ safety. Finally, self-reported adherence to COVID-19 public health measures and perceptions of susceptibility to and severity of COVID-19 were not associated with vaccine acceptance.

**Table 1.** Key characteristics, beliefs, knowledge and medical history and their associations with COVID-19 vaccine acceptance.

|  | **Total**<br>**(n=1037)** | **Acceptance of**<br>**COVID-19 vaccine**<br>**(n=684 (66.0%))** | **Refusal of COVID-**<br>**19 vaccine**<br>**(n=299 (28.8%))** | **Undecided**<br>**(n=54 (5.2%))** | **Refusal of**<br>**COVID-19**<br>**vaccine<sup>a</sup>** |
| --- | --- | --- | --- | --- | --- |
|  | **n (%)** | **n (%)** | **n (%)** | **n (%)** | **OR (95% CI)** |
| **Socio- Demographics** |  |  |  |  |  |
| **Age (years)** |  |  |  |  |  |
| 50-59 | 629 (60.7) | 420 (66.8) | 184 (29.2) | 25 (4.0) | 1 |
| 60-69 | 282 (27.2) | 184 (65.2) | 77 (27.3) | 21 (7.5) | 0.96 (0.70-1.31) |
| 70+ | 126 (12.2) | 80 (63.5) | 38 (30.2) | 8 (6.3) | 1.08 (0.71-1.66) |
| **Gender** |  |  |  |  |  |
| Male | 579 (55.8) | 394 (68.1) | 157 (27.1) | 28 (4.8) | 1 |
| Female | 458 (44.2) | 290 (63.3) | 142 (31.0) | 26 (5.7) | 1.23 (0.94-1.61) |
| **Residence** |  |  |  |  |  |
| Inside ITS | 402 (38.8) | 283 (70.4) | 102(25.4) | 17 (4.2) | 1 |
| Outside ITS | 635 (61.2) | 401 (63.2) | 197 (31.0) | 37 (5.8) | **1.36 (1.03-1.81)** |
| **Education** |  |  |  |  |  |
| Never attended school | 479 (46.4) | 317 (66.2) | 129 (26.9) | 33 (6.9) | 1 |
| Attended school | 554 (53.6) | 364 (65.7) | 169 (30.5) | 21 (3.8) | 1.14 (0.87-1.50) |
| **Health** |  |  |  |  |  |
| **Chronic conditions** |  |  |  |  |  |
| None | 277 (27.8) | 182 (65.7) | 80 (28.9) | 15 (5.4) | 1 |
| At least one | 718 (72.2) | 472 (65.7) | 207 (28.8) | 39 (5.4) | 1.00 (0.73-1.36) |
| **Hypertension** |  |  |  |  |  |
| No | 601 (60.6) | 394 (65.5) | 173 (28.8) | 34 (5.7) | 1 |
| Yes | 390 (39.4) | 258 (66.2) | 112 (28.7) | 20 (5.1) | 0.99 (0.74-1.31) |
| **Diabetes** |  |  |  |  |  |
| No | 771 (77.6) | 513 (66.5) | 220 (28.5) | 38 (4.9) | 1 |
| Yes | 222 (22.4) | 140 (63.1) | 66 (29.7) | 16 (7.2) | 1.10 (0.79-1.53) |
| **Vaccination Perceptions** |  |  |  |  |  |
| **I think vaccines are safe** |  |  |  |  |  |
| Agree | 670 (64.6) | 504 (75.2) | 144 (21.5) | 22 (3.3) | 1 |
| Neither agree nor disagree or Don't know | 211 (20.4) | 129 (61.1) | 68 (32.2) | 14 (6.6) | **1.84 (1.30-2.61)** |
| Disagree | 156 (15.0) | 51 (32.7) | 87 (55.8) | 18 (11.5) | **5.97 (4.03-8.84)** |
| **I think vaccines are effective** |  |  |  |  |  |
| Agree | 680 (65.6) | 510 (75.0) | 148 (21.8) | 22 (3.2) | 1 |
| Neither agree nor disagree or Don't know | 226 (21.8) | 135 (59.7) | 74 (32.7) | 17 (7.5) | **1.89 (1.35-2.65)** |
| Disagree | 131 (12.6) | 39 (29.8) | 77 (58.8) | 15 (11.4) | **6.80 (4.44-10.42)** |
| **COVID-19 Measure Adherence** |  |  |  |  |  |
| **Attended social events** |  |  |  |  |  |
| No | 934 (94.0) | 613 (65.6) | 269 (28.8) | 52 (5.6) | 1 |
| Yes | 60 (6.0) | 40 (66.7) | 18 (30.0) | 2 (3.3) | 1.02 (0.58-1.82) |
| **Mainly stayed at home except for essential purchasing** |  |  |  |  |  |
| No | 197 (19.8) | 127 (64.5) | 60 (30.4) | 10 (5.1) | 1 |
| Yes | 798 (80.2) | 527 (66.0) | 227 (28.5) | 44 (5.5) | 0.91 (0.65-1.29) |
| **Received visitors at home** |  |  |  |  |  |
| No | 675 (67.8) | 436 (64.6) | 197 (29.2) | 42 (6.2) | 1 |
| Yes | 320 (32.2) | 218 (68.1) | 90 (28.1) | 12 (3.8) | 0.91 (0.68-1.23) |
| **Worn a mask** |  |  |  |  |  |
| No | 128 (12.9) | 88 (68.8) | 36 (28.1) | 4 (3.1) | 1 |
| Yes | 866 (87.1) | 566 (65.4) | 250 (28.8) | 50 (5.8) | 1.08 (0.71-1.64) |
| **COVID-19 Perceived severity** |  |  |  |  |  |
| **COVID-19 is a serious infection that is spreading across the world** |  |  |  |  |  |
| True | 965 (95.9) | 642 (66.5) | 275 (28.5) | 48 (5.0) | 1 |
| False | 41 (4.1) | 24 (58.5) | 15 (36.6) | 2 (4.9) | 1.46 (0.75-2.82) |
| **COVID-19 Perceived susceptibility** |  |  |  |  |  |
| No | 228 (23.5) | 140 (61.4) | 75 (32.9) | 13 (5.7) | 1 |
| Yes | 741 (76.5) | 496 (66.9) | 206 (27.8) | 39 (5.3) | 0.77 (0.56-1.07) |

<sup>a</sup>The unadjusted ORs are for refusal of COVID-19 vaccine versus acceptance; the "undecided" were excluded from logistic regression analyses.

## Conclusion

These preliminary findings highlight important directions towards enhancing vaccine acceptance among Syrian refugees in Lebanon to reach herd immunity and ensure that Syrian refugees, who make up a significant proportion of the resident population in the country, are not left behind. In Lebanon, where Syrian refugees experience security, mobility, and healthcare access barriers, humanitarian organisations ought to consider delivery of immunization services to older refugees inside and outside ITSs. Practical approaches include disseminating accurate, accessible, and culturally appropriate information about vaccine safety and effectiveness. Importantly, these public health information campaigns must tackle the unprecedented surge of misinformation ‘infodemic’ about COVID-19 vaccines^9^; consider the integration of the Complacency, Convenience and Confidence (“3Cs”) model of vaccine hesitancy^10^; and work with community leaders, organisations and other key influencers to improve confidence, re-establish trust and address possible determinants influencing vaccine acceptability among Syrian refugees.

## Data Availability

The data that support the findings of this study are not publicly available but can be requested from the corresponding author for consideration for data sharing by the study team.

## Contributors

SA, HG, SM, SB and ZEK were involved in the conceptualization of the study and the survey design. SA and SM contributed to the literature search, interpretation of the data, and the writing of the manuscript. MH and NS contributed to data collection and analysis. NS contributed to the literature search and wrote the first draft of the paper. The underlying survey data were verified by SM and NS. All authors have reviewed, drafted and approved the final submitted version of the manuscript.

